# Predictive Modelling’s role in Improving Pre-exposure Prophylaxis (PrEP) Uptake in High-Risk HIV Groups in Africa: An Integrative Scoping Review

**DOI:** 10.1101/2025.09.20.25336235

**Authors:** Martin Zang Pam, Alex Odoom, Marian Serbeh

**Affiliations:** Department of Epidemiology and Disease Control, School of Public Health, University of Ghana, Legon, Accra Ghana; Department of Animal Health, Federal College of Animal Health and Production Technology, National Veterinary Research Institute, Vom, Plateau state, Nigeria; Department of Medical Microbiology, University of Ghana Medical School, Korle Bu, Accra, Ghana; Department of Social and behavioral science, School of Public Health, University of Ghana, Legon, Accra Ghana

**Keywords:** Predictive modelling, Preexposure prophylaxis (PrEP), High Risk Groups, Community-based strategies, Africa

## Abstract

This scoping review explores how predictive modelling can strengthen pre-exposure prophylaxis (PrEP) uptake among high-risk populations in Africa, where HIV prevalence remains disproportionately high. Although PrEP is highly effective (40–90%), its uptake and adherence remain suboptimal in LMICs. Predictive modelling provides a promising solution by identifying individuals at elevated risk, enabling targeted, evidence-based interventions. Using Arksey and O’Malley’s framework and PRISMA-ScR strategy, PubMed, Cochrane Library, ProQuest, and Google Scholar were searched for Africa-based studies from 2015–2025. Eligible studies focused on high-risk groups, including men who have sex with men, sex workers, persons who inject drugs, adolescents, and serodiscordant couples, and applied machine learning, regression models, deep learning, and neural networks.

Out of 209 records screened, 10 studies met inclusion criteria. Conducted between 2019–2025, they demonstrated how predictive tools can stratify HIV risk, enhance adherence monitoring, and improve resource allocation. Sixty percent relied on demographic and behavioural data and achieved strong predictive performance, particularly for HIV status prediction (70%). However, stigma, weak health systems, poor integration, and limited data quality still hinder implementation.

The review underscores predictive modelling’s transformative potential to scale PrEP services across Africa. Integrating machine learning, behavioural modelling, and community-based approaches can improve programmatic efficiency, equity, and targeting. Yet substantial gaps persist in translating predictive outputs into actionable interventions, addressing ethical issues, and validating models in diverse, resource-limited settings. Strengthening collaborations between data scientists, healthcare workers, and policymakers are essential to deliver cost-effective, context-specific PrEP services and accelerate HIV prevention efforts across the continent.

**KEY MESSAGES**

- **What is already known:** The use of Predictive modelling for identifying high-risk individuals to improve PrEP targeting, holds substantial promise for reducing HIV incidence among vulnerable groups, yet its integration into African health systems remains constrained by structural, data, and equity barriers.
- **What this study adds**: This scoping review demonstrates, for the first time, how diverse predictive modelling approaches like machine learning, deep learning, and clustering applied to epidemiological and behavioural data can enhance PrEP uptake and adherence among high-risk groups in LMIC African settings.
- **How this study could affect research, practice or policy:** The review findings highlight priority areas for integrating predictive tools with youth-friendly, community-based, and health system–strengthening strategies to scale cost-effective PrEP delivery, improve adherence, and guide evidence-based HIV prevention policy in Africa.

## Introduction

Since its discovery in the early 1980s, the human immunodeficiency virus (HIV) has remained one of the world’s foremost public health challenges, affecting approximately 38.4 million people globally. Despite significant advances in diagnosis, treatment and prevention, above 2.5 million new cases occur each year, mostly among people aged 15–49 years, with an estimated 19% unaware of their status.^1^ ^2^ Although global strategies aim to end the epidemic by 2030, persistent new infections, ongoing HIV-related comorbidities such as Tuberculosis, Oesophageal Candidiasis and Pneumocystis pneumonia—and health system limitations continue to hinder progress.^3^ These challenges are most pronounced in low- and middle-income countries (LMICs), especially across Africa, which carries the largest burden of HIV due to fragile healthcare systems, poverty, limited access to services, and overburdened infrastructures. Social stigma, harmful cultural norms, low education and widespread inequality further exacerbate the epidemic, complicating prevention and long-term treatment outcomes.^4–6^

Pre-exposure prophylaxis (PrEP) has recently come up as a cornerstone HIV-preventive strategy when taken consistently and as prescribed. It is particularly vital for high-risk populations (HRPs) such as men who engage in sex with men (MSM), persons who inject drugs (PWID), female sex workers (FSWs) and persons in serodiscordant relationships.^7–10^ Recognised by the World Health Organization as an essential intervention, PrEP’s success depends heavily on adherence, access to quality health services and public awareness. A comprehensive approach encompassing risk identification, monitoring, management of potential complications and adherence support is necessary to maximise PrEP’s impact.^11^ Scaling up PrEP across sub-Saharan Africa, which accounts for about 70% of the global HIV burden, could dramatically reduce new infections and bolster progress towards achieving the UNAIDS “Fast-track” 95–95– 95 targets.^12^ ^13^ These targets form the foundation for accelerating reductions in HIV incidence and moving towards the ultimate goal of ending the epidemic by 2030.

Predictive modelling has become an increasingly valuable tool in the control of infectious diseases, particularly in LMICs. By analysing disease trends, population movements and environmental factors, predictive models support early detection, outbreak tracking and timely interventions.^14^ In the context of HIV prevention, predictive modelling enhances PrEP uptake by identifying individuals most at risk, predicting adherence patterns and enabling more efficient outreach, personalised care and resource allocation.^15^ These tools are especially critical in resource-constrained settings where efficiency, timely action and minimising missed prevention opportunities are paramount.^14^ ^16^

Accurate estimation of HIV risk is a critical step in guiding testing campaigns, initiating PrEP and deploying targeted prevention strategies. While traditional models have been useful in identifying population-level risk, growing interest now centres on predictive modelling approaches that can refine individual- and community-level outcomes. Models such as Susceptible–Exposed–Infected–Recovered (SEIR) or Susceptible–Infected–Refractory– Susceptible (SIRS), as well as geospatial frameworks, have proven instrumental in epidemic forecasting and mapping high-prevalence areas, particularly in resource-inadequate locations. Agent-based models (ABMs) simulate behaviour-driven transmission dynamics, while machine learning (ML) and Markov models provide personalised risk prediction and cost-effectiveness analysis. Artificial intelligence (AI) is increasingly integrated to enable real-time surveillance and dynamic risk profiling.

This review therefore seeks to map and critically evaluate the application of predictive modelling tools in improving PrEP uptake and HIV prevention approaches among high-risk groups across the African continent. By synthesising existing evidence, it aims to identify knowledge gaps, highlight best practices and offer insights for more targeted, cost-effective and equitable PrEP delivery in low-resource settings.

### Review question

How can predictive modelling approaches be applied to identify high-risk populations and improve PrEP uptake and adherence among HIV-vulnerable groups in Africa?

### Review Objectives

To examine how predictive modelling identifies high-risk HIV groups and guides targeted PrEP strategies in Africa, and to assess the effectiveness of commonly used predictive models in improving PrEP uptake and adherence among vulnerable populations.

## Methods

The scoping review used the proposed methodology of Arksey and O’Malle (2005);^17^ addition to the scoping review types.^18^ It was put down in consonance with the Preferred Reporting Items in Systematic Reviews and Meta-analyses (PRISMA)

### Sources of data and search strategies

Data for this integrative scoping review were obtained from a comprehensive search of peer-revised and grey works, comprising primary research studies, systematic and narrative reviews, theoretical articles, editorials and relevant policy documents. Medical subject headings (MeSH) with relevant keywords were employed to design the investigation strategy derived from the study’s objectives and research questions. Core terms included: *HIV*, *Pre-exposure Prophylaxis (PrEP)*, *Predictive Modelling*, *High-Risk Populations*, and *Low-resource settings (Africa)*. Boolean operators (OR, AND), truncation, and phrase searching were used to refine and combine the search terms. Literature checks were performed across four major free-access electronic databases, including PubMed, Cochrane Library, ProQuest and direct Google

Scholar search, covering articles published between 2015 and 2025 (Table A.1 Appendix). All retrieved studies were first scrutinised from a downloaded CSV file while checking their title and abstract’s relatedness to the work, with full-text reviews performed for those meeting the study’s eligibility criteria. This rigorous approach ensured a broad yet focused synthesis of the existing knowledge on the topic.

### Study Selection

This review focused on reviewed materials such as article publications, book chapters, screened thesis works and policy documents from the last 10 years (2015–2025) that employed predictive modelling for HIV PrEP uptake across the African continent. Eligible studies included both quantitative and qualitative research involving adolescent/adult high-risk populations aged 18 and above—specifically female sex workers (FSWs), men engaging in sex with men (MSM), people who inject drugs (PWID), adolescents–young adults and serodiscordant cohorts. Only English-language studies that applied predictive modelling to improve PrEP uptake or adherence among these groups in the African setting were included. Studies published before 2015, conducted outside LMICs (Africa), or involving other key populations not specified above were excluded. Also excluded were articles lacking a predictive modelling component or those unrelated to HIV prevention. To enhance the breadth of the review, reference list mining and citation tracking were also conducted using Google Scholar to identify additional relevant studies. Two independent reviewers screened the titles and abstracts of the identified articles, and any inconsistencies were resolved via consultations with a third evaluator.

**Table 1.** Characteristics of the selected materials used in the review following the Population, Intervention Comparison and Outcome (PICO) format.

| Parameter | Description of the criteria used | Exclusion Description |
| --- | --- | --- |
| **Population (P)** | High-risk HIV populations across the African continent specifically:<br>– Men that have sex with men (MSM)<br>– Female sex workers (FSWs)<br>– People who inject drugs (PWID)<br>– HIV-serodiscordant couples and<br>– Adolescents and young persons in Africa | Research involving populations outside LMICs (e.g., high-income countries) or those not focused on key high-risk groups such as MSM, FSWs, PWID, or serodiscordant couples (e.g., general adult populations without defined HIV risk). |
| **Intervention (I)** | Application of predictive modelling techniques (e.g., machine learning, deep learning, AI-based risk algorithms, and statistical forecasting tools) to the following:<br>– Identify high-risk individuals<br>– Predicting the HIV infection risk<br>– Improved pre-exposure prophylaxis (PrEP) uptake and adherence | Studies that do not involve predictive modelling, machine learning, or data-driven tools for HIV prevention (e.g., only behavioural or pharmaceutical interventions without a data prediction component). |
| **Comparator (C)** | Standard or conventional approaches such as<br>– Non-predictive, population-wide risk assessments<br>– General PrEP delivery strategies without personalised targeting<br>– Traditional statistical or surveillance methods (e.g., static models) | Research lacking a comparative element, such as those that do not contrast predictive models with standard approaches or do not assess the impact against any benchmark. |
| **Outcome (O)** | Improvements in<br>Identification of high-risk individuals<br>– PrEP uptake<br>– Adherence and retention<br>– Cost-effectiveness of PrEP programmes<br>– Policy guidance for efficient HIV prevention in Africa | Studies that do not report on PrEP uptake, adherence, retention, or risk prediction outcomes or fail to connect predictive models with meaningful HIV prevention outcomes across Africa. |

### Data extraction

The data extraction began with a download of a CSV file and was refined using a Microsoft Excel (365) spreadsheet. Pooled studies were outlined on a pre-planned data retrieval pattern with features like study author(s), title, abstract, year study was conducted, key study take away/findings and country where the work was conducted in Africa recorded and summarized in a pre-formed table.

### Data synthesis approach/Analysis

This scoping review applied the Arksey and O’Malley outline, complemented by Joanna Briggs Institute (JBI) methodology, to systematically identify and synthesize evidence on predictive modelling for improving PrEP uptake among high-risk HIV groups in Africa. Data were thematically organized by target populations, study context and PrEP outcomes, publication year, predictive model types, and study locations. Quantitative findings like initiation and adherence rates, PrEP efficacy measures, and model performance were summarized descriptively in tables and figures. Qualitative data on implementation barriers, facilitators, and contextual factors underwent thematic analysis to identify patterns, gaps, and opportunities for scaling predictive approaches. This integrative process enabled a comparative understanding of how predictive tools operate across diverse African settings, their implications for equity and scalability, and the contextual conditions needed for effective implementation. By combining quantitative and qualitative evidence, the review highlights benefits, limitations, and key research gaps to inform future applications in LMICs.

### Consent and ethical considerations

The study did not directly involve participants, stakeholders or communities, hence consent or ethical clearance was not applicable.

## Results

### Search result

A total of 209 records were recovered from PubMed, Cochrane Library, ProQuest, and Google Scholar, covering studies from 2015 to 2025. After excluding 135 irrelevant records and removing 4 duplicates, 70 articles remained. Fifty-five (55) studies were screened out for not using predictive modelling, leaving 15 that met the study’s partial eligibility. After the full text review, 5 studies were removed because they were not done in an LMIC (African setting), did not include high-risk populations, or misaligned with the review objectives, leaving 10 eligible works that fully met the studies objectives and inclusion criteria (Figure 1. below).

**Figure 1.**
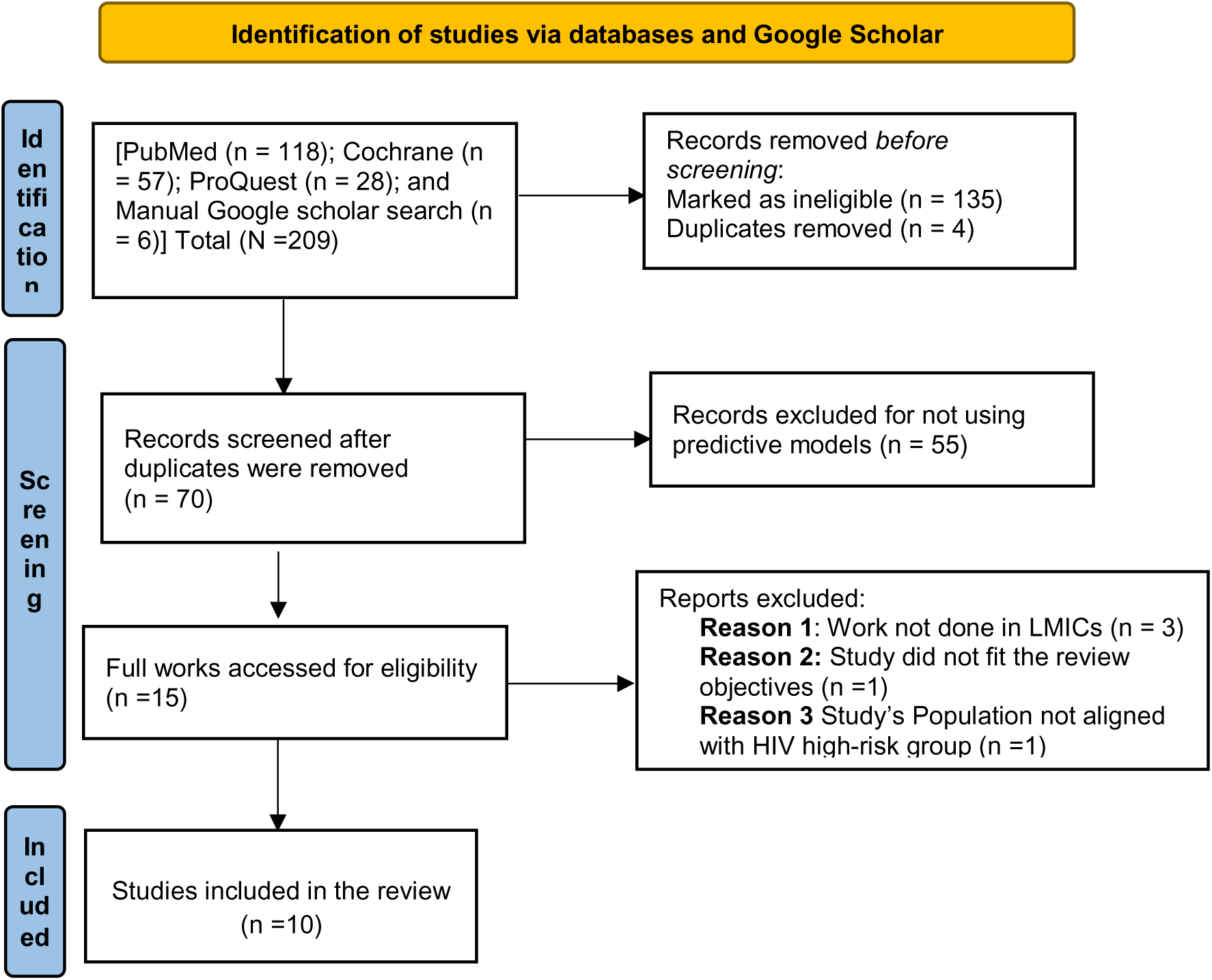
PRISMA Flow chart of the screened studies used in the scoping review

### Characteristics of the included studies

Table 2 summarises the 10 studies that met the inclusion criteria for this scoping review, all conducted between 2019 and 2025 in various African countries. These studies collectively investigated the application of predictive modelling tools to improve the identification of individuals at elevated risk of contracting HIV, support PrEP uptake, and enhance adherence across diverse vulnerable populations. The populations targeted in these studies were female sex workers (FSWs), men who have sex with men (MSM), people who inject drugs (PWID), HIV-serodiscordant couples, and in some cases, general high-risk adolescents and youth. The geographical coverage of the studies spans countries such as Zimbabwe, South Africa, Ethiopia, Uganda, Rwanda, and Kenya and included multi-country analyses across Eastern and Southern Africa, reflecting regional diversity in data, risk profiles, and healthcare system challenges.

**Table 2.** Summary of the study characteristics.

| Author(s) | Title | Study year | Key Take away | Source/Study type | Predictive model used | Study Population and Location in Africa |
| --- | --- | --- | --- | --- | --- | --- |
| Hunidzarira et al.<sup>19</sup> | Sexually Transmitted Infections and Human Immunodeficiency Virus (HIV)-1 Among African Women | 2022 | Sexually transmitted infection incidence did not reliably predict HIV-1 incidence at the population level among at-risk African women partaking in 2 large PrEP trials. | Journal of Infectious Diseases/Rand omised trial | Regression modelling | Women from 4 African countries (4 countries (Malawi, South Africa, Uganda, and Zimbabwe) |
|  | Participating in HIV-1 Pre-Exposure Prophylaxis Trials |  |  |  |  |  |
| Siyamayambo et al.<sup>20</sup> | Diagnosis and Management of Sexually Transmitted Infections Using Artificial Intelligence Applications Among Key and General Populations in Sub-Saharan Africa: A Systematic Review and Meta-Analysis | 2025 | Artificial intelligence and machine learning applications show high potential in improving the diagnosis, treatment, and management of sexually contracted infections such as HIV in Sub-Saharan Africa, especially among key populations, although their real-world implementation remains limited. | Algorithms/Systematic review and meta-analysis | Fourth Industrial Revolution (4IR) technologies being artificial intelligence (AI) and machine learning (ML-random forest and XGBoost | Not specifically mentioned and conducted from 20 applicable studies in 20 African countries, including South Africa, Ethiopia and Zimbabwe |
| Grant Hannah<sup>21</sup> | The scale-up of PrEP for HIV prevention in high-risk women in sub-Saharan Africa: use of mathematical modelling to inform policy making | 2020 | PrEP remains the most cost-effective intervention for HRGs such as female sex workers as expanding access can reduce HIV infections broadly. Effective scale-up requires balancing cost with wide integration into health systems. Static models aid short-term planning, whereas dynamic models suit evolving epidemics and long-term policy decisions. | London School of Hygiene and Tropical Medicine/Doctoral dissertation paper | Mathematical Modelling (static and dynamic models) | Women (sex workers) across SSA |
| Belete, and Manjaiah.<sup>22</sup> | A Deep Learning Approaches for Modelling and Predicting HIV Test Results Using the EDHS Dataset | 2023 | The study demonstrates that deep learning models applied to Ethiopian demographic and health survey data can accurately predict individuals' HIV test status, offering a valuable tool for informing early intervention strategies and policymaking | Intech Open/Retrospective study using secondary data | The deep learning model | Not specifically mentioned and Ethiopia (using the country's demographic health survey (EDHS) datasets |
| Birri-Makota and Eustasius.<sup>23</sup> | Predicting HIV infection in the decade (2005–2015) pre-COVID-19 in Zimbabwe: A supervised classification-based machine learning approach | 2023 | The work identified machine learning models, particularly XGBoost, to accurately predict HIV infection risk in Zimbabwe using demographic and behavioural data, with total lifetime sexual partners and cohabitation duration emerging as key predictors for females and males, respectively. | PLOS Digital Health/Retrospective study using secondary datasets | Machine learning model (XGBoost) | Males and Females in Zimbabwe |
| Chingombe et al.<sup>24</sup> | Predicting HIV status among men who have sex with men in Bulawayo & Harare, Zimbabwe using bio-behavioural data, recurrent neural networks and machine learning techniques | 2022 | The study showed that recurrent neural networks (RNNs) accurately forecast HIV status among MSM in Zimbabwe, outperforming other machine learning models and offering a powerful tool to enhance early HIV screening and service delivery for this marginalised population | Tropical Medicine and Infection Diseases/Retro spective cross-sectional study | Recurrent neural networks and machine learning | Men who engage in sex with other men in Zimbabwe |
| Oladokun, O. O.<sup>25</sup> | Predicting the HIV status among women in South Africa using machine learning: comparing the decision tree model and logistic regression | 2019 | The study finds that logistic regression slightly outperforms decision tree models in forecasting HIV status among women in South Africa via the use of demographic data. However, both models show limited accuracy, suggesting that demographic variables alone may be insufficient for reliable HIV prediction | Master's Thesis submitted to the University of the Witwatersrand, Johannesburg (South Africa)/Retros pective cross-sectional study | Machine learning (comparing logistic regression to decision tree models) | Women in South Africa |
| Orel et al.<sup>26</sup> | Machine Learning to Identify the Socio-Behavioural Predictors of HIV Positivity in East and Southern Africa | 2020 | Findings from this research reveal that XGBoost effectively predicts HIV status using socio-behavioural data from 10 East and Southern African countries, enabling the identification of people living with HIV and high-risk individuals for targeted testing and prevention strategies such as PrEP and male circumcision. | MedRxiv/Retr ospective Quantitative cross-sectional study using secondary data analysis from national surveys | Four machine-learning algorithms including XGBoost | Males and Females in 10 East and Southern African countries |
| Mutai et al.<sup>27</sup> | Use of unsupervised machine learning to characterise HIV predictors in sub-Saharan Africa | 2023 | The study demonstrates that unsupervised machine learning specifically hierarchical clustering effectively identifies groups of SSA countries with similar socio-behavioural HIV risk profiles, highlighting key predictors such as education, circumcision and urban residence for targeted screening and intervention strategies. | BMC Infectious Diseases/Retro spective exploratory cross-sectional study | Machine learning (an unsupervised agglomerative hierarchical model) | Males and Females from 13 Sub-Saharan African Countries |
| Balzer et al.<sup>28</sup> | Machine learning to identify persons at high risk of human immunodeficiency virus acquisition in rural Kenya and Uganda | 2019 | Using >75% population-level HIV testing data in rural Kenya and Uganda, machine learning (Super Learner) outperformed the risk-group and model-based approaches in identifying individuals at high risk for HIV acquisition. It improves both efficacy and sensitivity by preventing HIV among the main groups in high-incidence settings. | Clinical Infectious Diseases/Longitudinal Population-Based Cohort Study | Logistic regression (based risk score) versus machine learning risk score (Super Learner algorithm) | Rural Kenyan and Ugandan communities |

A wide array of predictive modelling approaches was employed across the studies. These included machine learning models (e.g., XGBoost, random forest, decision trees), deep learning models (such as recurrent neural networks, RNNs), regression-based models, and unsupervised learning techniques such as agglomerative hierarchical clustering. These tools were applied to analyse large-scale datasets derived from national demographic and health surveys to detect patterns in socio-behavioural, demographic, and clinical variables that are predictive of HIV status or future risk. The study designs varied widely, encompassing retrospective cross-sectional studies, quantitative data analyses, systematic reviews, meta-analyses, and doctoral research projects. Although some studies focused on model comparison to assess predictive accuracy, others explored implementation strategies, cost-effectiveness, and population-level impact, offering a comprehensive picture of how predictive models can guide targeted PrEP interventions in resource-limited settings (Table 2).

### Thematic Categorisation of included studies

Thematic and key findings from the included studies were grouped into categories (model types, geographic relevance, target populations, predictor variables, and outcome relevance).

**Table 3:** Thematic Categorisation and Frequency of Key Findings Across 10 Studies on Predictive Modelling and PrEP-Related HIV Risk in African LMICs.

| Thematic Category | Description | Frequency (N=10) | Examples |
| --- | --- | --- | --- |
| *Use of ML/AI Tools* | Application of AI/ML models such as XGBoost, RNNs, deep learning, clustering | 10/10 | XGBoost for risk prediction, RNNs for MSM HIV status, hierarchical clustering for SSA countries |
| *HIV status prediction focus* | Models developed to predict HIV test outcomes or risk levels | 7/10 | Ethiopia DHS data, MSM-focused screening tools, Zimbabwe risk models |
| *Key predictors identified* | Behavioural/demographic risk variables found to be important in prediction models | 5/10 | Lifetime sexual partners, cohabitation, education, and urban residence |
| *Application to PrEP and Targeted Interventions* | Use of findings to inform PrEP targeting, male circumcision, or testing | 3/10 | PrEP for FSWs, risk-based HIV screening |
| *Focus on Key Populations (HRGs)* | Inclusion of MSM, FSWs, AGYW, PWID, or serodiscordant couples | 4/10 | MSM in Zimbabwe, FSWs for PrEP rollout, AGYW risk profiling |
| ***Model Comparisons and Benchmarking*** | Studies comparing ML methods (e.g., logistic vs. tree models) | 4/10 | Logistic regression is decision trees in South Africa |
| ***Use of Socio-Behavioural Data*** | Reliance on demographic/behavioural data for modelling | 6/10 | DHS data from 10 countries and survey-based behavioural profiles |
| ***Policy and Health System Relevance*** | Models linked to policy use or system planning | 5/10 | Dynamic/static models for planning and national risk clustering |
| ***Limitations Identified*** | Gaps in model accuracy, lack of STI-HIV correlation, or real-world deployment | 2/10 | STI not predictive of HIV, limited AI real-world integration |

### Pictoral themes from the 10 included study findings

The frequency of thematic findings across the 10 studies on predictive modelling for HIV prevention in African LMICs (Figure 2) revealed the dominance of AI/ML applications, key populations focus, socio-behavioural data use, and implications for PrEP delivery and health policy

**Figure 2.**
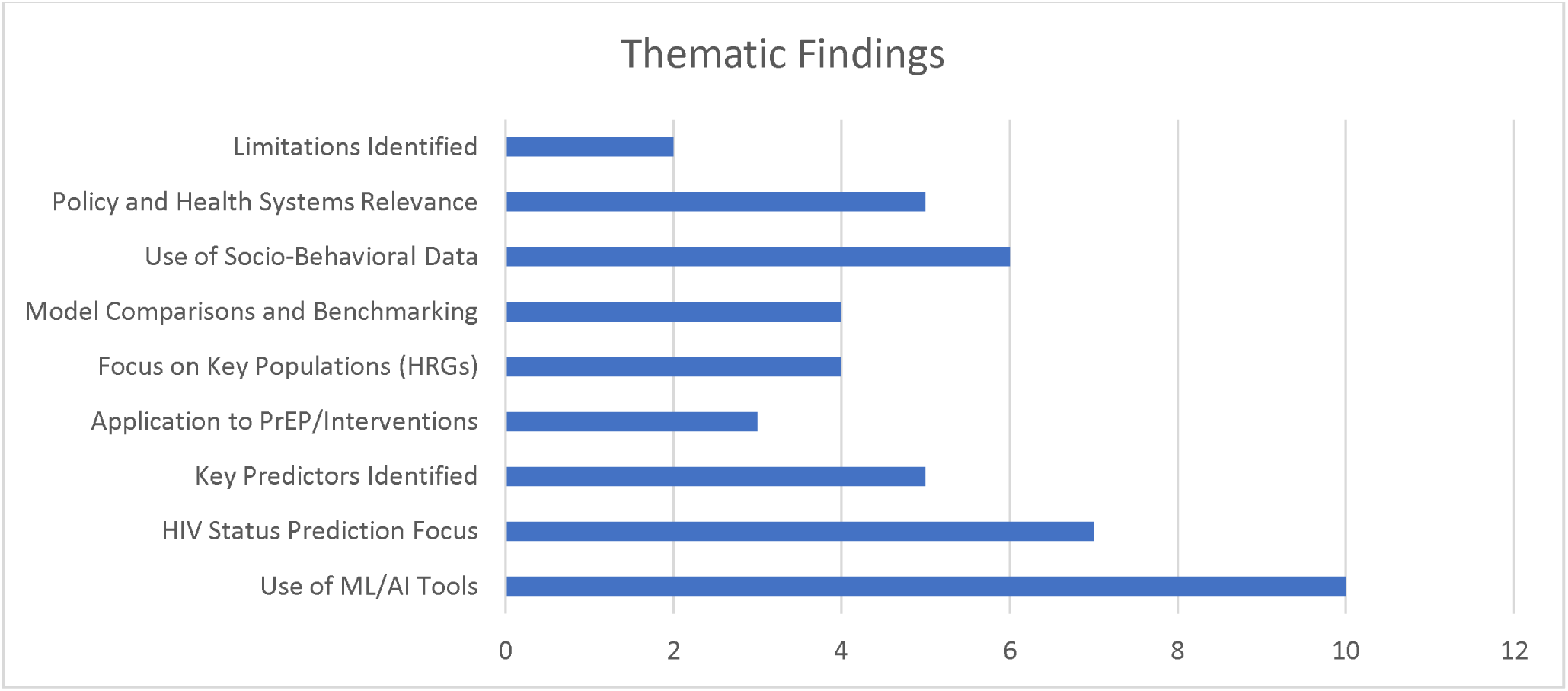
Chart showing the thematic frequency of findings across the 10 studies on predictive modelling for HIV prevention in African LMICs

## Discussion

Predictive models improve the identification of high-risk individuals and can increase pre-exposure prophylaxis (PrEP) uptake, but successful implementation in Africa and other LMICs requires addressing contextual barriers, fostering key facilitators aimed at increasing PrEP implementation outcomes and ensuring effective risk communication. Predictive modelling has emerged as a valuable tool for improving PrEP uptake among high-risk HIV groups in Africa by enabling more precise identification of individuals at greatest risk and informing targeted interventions.

This scoping review examined 10 studies^19–28^ conducted across some African countries between 2019 and 2025. These studies applied various predictive modelling techniques such as machine learning (ML), logistic regression, decision trees, deep learning, and neural networks to enhance PrEP acceptance among high-risk HIV individuals. The models assessed demographic, behavioural, clinical, and geographic data to identify individuals at elevated HIV risk from the key populations studied. Some of the predictive accuracies reported from these studies were in line with those seen from similar works reported by Rosenberg et al., and Romero et al., ^29^ ^30^ who described same especially when incorporating variables like sexual partner behaviours, HIV viremia, and local prevalence rates. For example, integrating predictive tools into clinical workflows in South Africa and Uganda significantly increased PrEP eligibility identification and prescriptions. Grant (2020)^21^ emphasised in her work the role such predictive data-driven tools play in enhancing cost-effective PrEP scale-up among high-risk populations, while Hunidzarira et al. ^19^ found that using sexually transmitted infection incidence alone was insufficient— highlighting the need for multifactorial models. Models observed using national health survey data in Ethiopia and supervised machine learning in Zimbabwe have showed promising results. ^22–24^ These models successfully pinpointed predictors such as lifetime sexual partners, cohabitation duration, and socio-behavioural factors emphasising the value of tailoring models to local epidemiological and social contexts.

Despite technical success, many studies acknowledged limited real-world impact due to structural and behavioural barriers. Challenges such as stigma, low risk perception, fear of side effects, and socio-economic constraints continue to impede PrEP uptake in.^31–33^ These findings suggest that predictive modelling must be paired with culturally sensitive, community-driven approaches, especially youth-friendly services, to translate risk identification into effective intervention. While predictive modelling offers powerful tools for targeting HIV prevention efforts, its success depends on aligning technological innovation with health system readiness and community engagement strategies.

### Implications for Implementation and Challenges in Policy, Practice, and Research

This review reveals critical implications for applying predictive modelling in real-world PrEP delivery across Africa and other LMICs. First, predictive analytics present a cost-effective strategy for prioritising limited HIV prevention resources, particularly in targeting interventions towards high-risk populations. This approach aligns with the WHO and UNAIDS recommendations that emphasise scaling up PrEP services in regions with high HIV incidence and low uptake, especially among adolescent girls and young women (AGYW), MSM and other main populations.^34^ ^35^

Secondly, integrating predictive tools into national HIV information systems and electronic health records (EHRs) could enhance real-time decision-making and enable data-driven resource allocation. However, implementation remains limited in LMICs due to infrastructural gaps, financial constraints, and a shortage of personnel trained in data science and artificial intelligence (AI). Addressing these barriers will require government investment in workforce development and strategic public-private partnerships to advance digital health ecosystems. In addition, ethical and regulatory frameworks must guide the predictive model deployment. These include ensuring a transparent model design, safeguarding patient data privacy, and preventing algorithmic bias that may intensify health inequities among already marginalised groups. Community engagement is vital for building trust, especially in settings with historical mistrust in healthcare systems.

From a research standpoint, future studies in the fields of implementation science should assess the level of predictive modelling usage and accuracy to examine the practical translation of such model outputs in improving PrEP initiation, retention, and adherence. Mixed-methods research that integrates implementation research can provide insights into the contextual effectiveness of predictive modelling whilst looking at gaps from the health systems that can impede the precipitation of better PrEP outcomes. Cost-effectiveness analyses and evaluations of model scalability and cross-border adaptability will also be critical for shaping sustainable, region-wide strategies.

Despite the demonstrated predictive accuracy, most models remain underutilised in routine clinical practice. Siyamayambo et al.^20^ noted the limited integration of Fourth Industrial Revolution (4IR) technologies such as machine learning and AI into African health systems, citing infrastructural and capacity challenges. Furthermore, many models rely on retrospective secondary data and underrepresent socio-behavioural factors such as stigma, disclosure fears, and perceived risk, limiting their community relevance. Some studies have showed that including variables such as education, circumcision, and urban residency improved model performance.^26^ ^27^ However, data standardisation across countries remains a major challenge. To bridge the gap between modelling and implementation, hybrid strategies are needed. Some advocates have suggested that predictive strategies should be embedded into culturally sensitive, youth-friendly, and community-led service platforms to ensure that risk identification leads to actionable and equitable PrEP delivery.^32^ ^33^

### Strategies Adopted to Enhance PrEP Initiation and Continuation and Lessons for Broader LMICs

The scoping review by Ekwunife et al.^36^ highlights strategies to improve pre-exposure prophylaxis (PrEP) initiation and continuation among adolescent girls and young women (AGYW) in LMICs. Recognising AGYW’s heightened HIV risk in Africa, the authors propose a three-pillar framework built around demand creation, PrEP initiation, and PrEP continuation.

### Demand Creation

To stimulate demand, social marketing campaigns and behaviour-centred design are emphasised. These efforts reframe PrEP as a tool of empowerment and protection rather than solely biomedical intervention. Peer influencer networks also play a key role by challenging stigma, providing relatable narratives, and communicating PrEP benefits via culturally relevant channels and youth-friendly platforms, including social media.

### PrEP Initiation

Common barriers such as limited knowledge, stigma, cost, and fear of side effects necessitate multifaceted strategies involving education, resource allocation, insurance coverage, and technology-driven solutions to increase accessibility.^37^ The importance of decision-support tools and adolescent-friendly health services to facilitate informed PrEP decisions, address adherence or side-effect concerns, and improve trust in prevention services is also stressed.^36^ AGYW are more likely to initiate PrEP in youth-focused or private clinics than in general family planning settings, highlighting the value of privacy, confidentiality, and nonjudgmental service environments. Establishing dedicated youth spaces or partnering with trusted community providers can further lower barriers.

### PrEP Continuation

Retention remains a major challenge due to AGYW’s rapidly changing life circumstances. Recommended interventions include SMS adherence reminders, drug-level feedback, peer-support clubs, and conditional economic incentives. Integrating psychosocial support and counselling into follow-up visits reinforces adherence and addresses stigma, mobility, and partner disclosure concerns. Differentiated service delivery (DSD) models such as community pharmacies, mobile outreach, and telemedicine bring services closer to where AGYW live and work. These decentralised approaches enhance privacy, flexibility, and convenience, particularly in rural or underserved areas.

### Lessons from Broader LMIC Contexts and Other Key Populations

While the focus of Ekwunife et al.^36^ is on AGYW, similar strategies have been successfully applied to other key populations at high risk of HIV in LMICs. For instance, studies from Kenya and South Africa have shown that peer-led education, mobile PrEP clinics, and community drop-in centres have significantly increased PrEP uptake and adherence amongst MSM’s and FSWs.^38^ ^39^ These interventions are often accompanied by stigma reduction campaigns and linkage to other services, such as STI screening and psychosocial support.

In Nigeria, key population-led clinics have emerged as a promising model for delivering PrEP to people who inject drugs (PWID) and HIV-serodiscordant couples. These clinics are typically staffed by members of the key populations themselves, fostering a non-judgmental and inclusive environment that encourages retention in care.^40^ In addition, digital tools have been widely adopted to support adherence and monitoring. In Uganda, the use of real-time adherence devices such as Wisepill and social media-based support platforms (e.g., WhatsApp groups) has shown promise in improving adherence among MSM. Meanwhile, in Zimbabwe and Zambia, community-based PrEP delivery through mobile health vans and integrated sexual and reproductive health (SRH)-HIV services has facilitated better outreach and engagement, particularly among underserved and mobile populations.^41^ These lessons highlight the need for population-specific tailored, flexible delivery mechanisms and strong community engagement to ensure that PrEP is not only accessible but also acceptable and sustainable.

### Suggested Strategies Using Boosters for PrEP Delivery in High-Risk Populations

#### Employing Implementation Research (IR)

Integrating implementation research (IR) into HIV prevention efforts can substantially improve pre-exposure prophylaxis (PrEP) delivery and effectiveness, particularly among high-risk groups (HRGs) in Africa and globally. This scoping review highlights the importance of adopting IR principles to address gaps in PrEP service delivery and uptake. Through methods such as literature reviews, stakeholder engagement, and contextual analyses, IR identifies key challenges including limited Demographic and Health Survey (DHS) data, stock management issues, and programmatic inefficiencies. Implementation strategies such as training healthcare professionals, supporting community health workers, and developing tailored educational programmes on demographic data use, digital tools, and predictive models can strengthen PrEP services.^36^ ^42–44^

When aligned with target groups’ needs, IR interventions have shown measurable improvements in PrEP initiation, adherence and retention. Predictive modelling can amplify these efforts in resource-limited settings by stratifying populations based on HIV risk and enabling efficient, locally tailored interventions while overcoming stigma, low awareness, and ethical barriers. Exploring both users’ and non-users’ perspectives enhances understanding of PrEP acceptability, feasibility, and service gaps.

Evidence from initiatives such as Australia’s EPIC-New South Wales—achieving a 25% HIV incidence reduction within one year, demonstrates the potential of high-coverage, targeted PrEP programmes.^45^ However, uptake and adherence remain suboptimal across many LMICs in Africa due to stigma, cost, poor integration, and limited provider engagement.^46^ ^47^ Community-based and telemedicine-supported PrEP programmes have proven feasible, acceptable, and scalable but require further adaptation to reach underrepresented groups, including women, people who inject drugs, and ethnic minorities.^43^ ^48^ Ultimately, IR especially when guided by predictive modelling offers a critical pathway for adapting HIV prevention strategies to local contexts, enhancing programme efficiency, and ensuring PrEP reaches those at highest risk, thereby maximising its public health impact.^45^ ^46^ ^48^

#### Role of Community Engagement

Community engagement through youth-friendly, culturally competent, and socially responsive approaches is a powerful strategy to strengthen PrEP delivery among high-risk HIV populations in Africa, including MSM, adolescences youth and adults, and female sex workers (FSWs). These methods reduce barriers such as stigma, discrimination, and distrust of health systems that often impede HIV prevention. Evidence shows that community-based interventions using non-judgmental communication and culturally sensitive messaging normalise PrEP use and increase confidence in prevention efforts, particularly in underserved or conservative settings.^49^ ^50^ Culturally relevant health education delivered by peer educators, community influencers, and digital platforms raises awareness, dispels myths, and supports informed decision-making around PrEP.^51^ Youth-friendly, confidential services and mobile outreach reduce access gaps caused by transportation or service limitations.^52^ ^53^ Direct community involvement in programme design fosters ownership, sustainability, and effectiveness. Models such as decentralised services, peer-led interventions, and integration with sexual and reproductive health services have further enhanced accessibility.^33^ ^54^ Engagement of trusted leaders, use of safe spaces, and youth-trained providers also improve uptake and adherence.^32^ ^55^ ^56^

Despite notable gains, challenges like stigma, side effects, limited resources, and community instability persist, requiring continuous adaptation, stakeholder engagement, and innovative demand-creation strategies. Expanding PrEP delivery to schools, youth zones, and community venues has yielded positive outcomes.^33^ ^54^ Ultimately, community engagement is central to scaling PrEP and reducing HIV incidence across Africa’s most vulnerable populations.

### Using the Health Systems Strengthening (HSS) Approach

Strengthening health systems is critical for effectively using predictive models to enhance HIV PrEP uptake among high-risk groups. Machine learning-based HIV risk prediction models integrated into electronic health records (EHRs) improve identification of PrEP-eligible individuals and prescription rates, often outperforming traditional risk assessments while maintaining equitable sensitivity across diverse populations.^30^ ^57–60^ Mathematical and simulation studies show that targeted PrEP delivery combined with HIV testing and behavioural interventions can substantially reduce new infections in high-burden settings.^61^ ^62^ Differentiated service delivery models such as peer-led and telemedicine-assisted programmes further improve PrEP adherence and retention.^43^ Successful implementation depends on integrating predictive tools into clinical workflows, training regimens, and addressing stigma, trust, and digital literacy concerns.^63^ Investment in digital infrastructure, provider capacity, and community engagement will enable ethical, precise, and scalable predictive analytics for PrEP delivery in resource-limited African contexts.^60^ ^58^ ^30^

**Figure 3.**
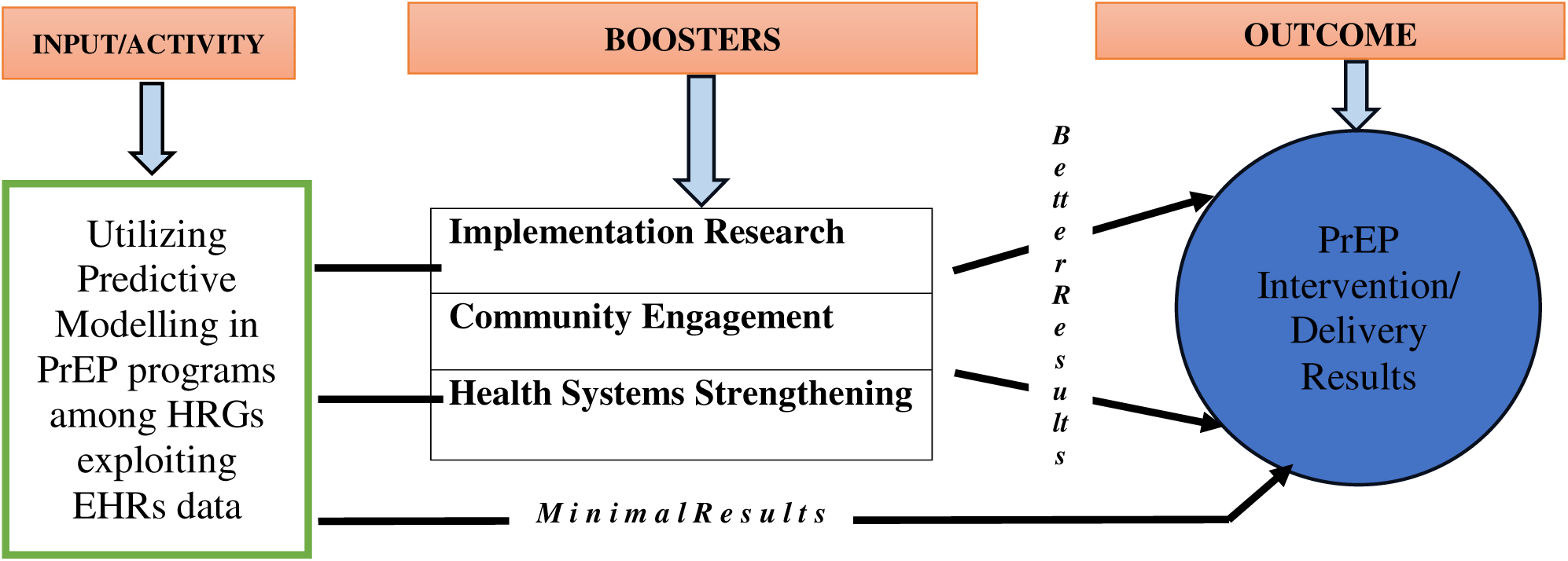
Suggested Framework for improving worldwide PrEP adoption using the 3 Booters approach across Africa

## Conclusion

Predictive modelling offers a powerful, data-driven approach to identify high-risk populations for HIV and guide targeted PrEP interventions in African LMICs. By integrating demographic, behavioural, clinical, and contextual data, these models can improve efficiency, cost-effectiveness, and scalability of HIV prevention. Machine learning approaches such as XGBoost, recurrent neural networks, and deep learning show strong potential for predicting HIV risk and informing early interventions. Complementary strategies, including implementation research, community engagement, and health systems strengthening, are essential to overcome structural and financial barriers, sustain adherence, and optimise programme impact. Tailoring predictive tools to key populations and socio-behavioural contexts enables more precise, equitable PrEP delivery, accelerating progress toward reduced HIV incidence and improved public health outcomes across the continent.

## Contributions

**MZP** conceptualized the study, database search and wrote the study’s first draft. **AO** also executed the study search and redrafting of the manuscript, while **MS** did the article’s final review and corrections

## Patient consent for publication

Not applicable

## Ethics approval

There was no human contact or need for primary records for this scoping review

## Data Availability

The required data for this work has been captured in the work

## APPENDICES

**Table A.1:**
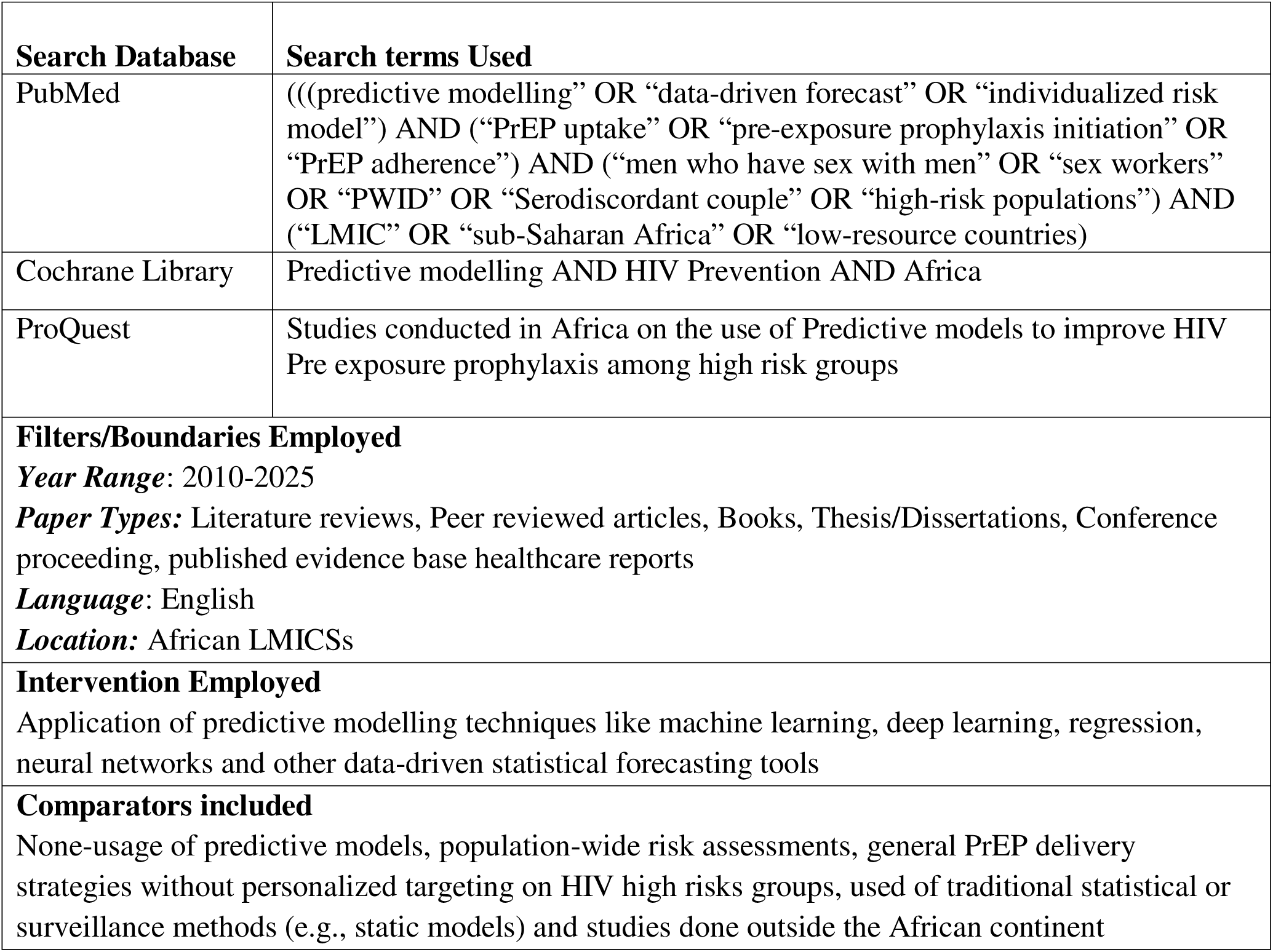
Databases and search terms for scoping review.

| **Search Database** | **Search terms Used** |
| --- | --- |
| PubMed | ((("predictive modelling" OR "data-driven forecast" OR "individualized risk model") AND ("PrEP uptake" OR "pre-exposure prophylaxis initiation" OR "PrEP adherence") AND ("men who have sex with men" OR "sex workers" OR "PWID" OR "Serodiscordant couple" OR "high-risk populations")) AND ("LMIC" OR "sub-Saharan Africa" OR "low-resource countries)) |
| Cochrane Library | Predictive modelling AND HIV Prevention AND Africa |
| ProQuest | Studies conducted in Africa on the use of Predictive models to improve HIV Pre exposure prophylaxis among high risk groups |
| **Filters/Boundaries Employed**<br>*Year Range:* 2010-2025<br>*Paper Types:* Literature reviews, Peer reviewed articles, Books, Thesis/Dissertations, Conference proceeding, published evidence base healthcare reports<br>*Language:* English<br>*Location:* African LMICs |  |
| **Intervention Employed**<br>Application of predictive modelling techniques like machine learning, deep learning, regression, neural networks and other data-driven statistical forecasting tools |  |
| **Comparators included**<br>None-usage of predictive models, population-wide risk assessments, general PrEP delivery strategies without personalized targeting on HIV high risks groups, used of traditional statistical or surveillance methods (e.g., static models) and studies done outside the African continent |  |

